# Preoperative multi-nutrient medical food versus fasting control in adults undergoing elective surgery: a single-center pilot trial

**DOI:** 10.64898/2026.02.12.26345765

**Authors:** Benjamin Zimmerman, Joshua Zvi Goldenberg, Thea Marx

## Abstract

**Background:** The surgical stress response is a predictable, physician-managed metabolic state triggered by anesthesia and tissue injury, marked by insulin resistance and hypercatabolism that create unique nutritional needs unmet by standard, pre-surgical fasting diets. We developed a multi-nutrient medical food to support perioperative metabolic homeostasis and piloted its safety/tolerability and exploratory outcomes.

**Methods:** In a single-center pilot trial (n=67) of adults undergoing elective abdominal, cardiac/thoracic, gynecological, or orthopedic surgery, participants were allocated to medical food or no-treatment control. The product was taken twice preoperatively (evening before and 4 h pre-op) with standard care. Primary safety outcomes were adverse events, postoperative nausea/vomiting (PONV), 30-day readmission, and infections. Exploratory outcomes were fasting glucose, HbA1c, electrolytes, cortisol, pre-operative emotional state, and post-operative pain.

**Results:** All participants completed the intervention. No product-attributed adverse events occurred. Gastric clearance was achieved within 2 h in all, and there were no 30-day readmissions or infections. PONV occurred in 30.3% vs 35.3% (risk ratio 0.86, 95% CI 0.43–1.71, *p*=0.796). Post-operative glycemia favored the intervention; at 48 hr the intervention group showed lower glucose (HL −9 mg/dL, *g*=0.35, *p*=0.030), while earlier timepoints were nonsignificant. Post-operative magnesium was numerically lower with intervention (4.76 vs 5.10) without statistical significance; other electrolytes and cortisol showed minimal differences. Post-operative pain was 5.33 vs 5.62 (*g*=0.19, *p*=0.43). Positive pre-operative emotion was more frequent with intervention (17/33 vs 9/34; risk ratio 1.95, *p*=0.046).

**Conclusion:** The medical food was safe and well tolerated without increased PONV or readmissions. Preliminary metabolic and emotional signals justify a larger, adequately powered efficacy trial.

**Clinical Relevancy Statement:** This pilot trial demonstrates that a preoperative multi-nutrient medical food was well tolerated and feasible to administer in a routine clinical setting: all participants achieved gastric clearance within 2 hours of the pre-operative dose, with no increase in PONV and no readmissions. Exploratory findings indicate potential benefits that could nutritionally support recovery if confirmed. These results support the feasibility of administering a targeted nutrition intervention shortly before surgery and justify evaluation in a larger efficacy trial.

**Clinical Trial Registration:** NCT07359222

## Introduction

The surgical stress response (SSR) is an acute, predictable, physician-managed metabolic state triggered by anesthesia and surgical tissue injury, characterized by insulin resistance, hypermetabolism, and catabolism, creating unique nutritional needs unmet by standard fasting diets and thus warranting controlled perioperative enteral nutrition.(1,2) SSR’s neuroendocrine–inflammatory cascade commonly leads to postoperative hyperglycemia and protein breakdown, both of which correlate with infection, organ dysfunction, and other complications.(3–6) Preoperative glycemic status also matters: elevated HbA1c has been associated with increased postoperative complications after major elective surgery, underscoring the value of metabolic optimization before and after the operative insult.(7)

Enhanced Recovery After Surgery (ERAS) pathways address the stress response with bundled measures, including abbreviated fasting and early oral intake.(8) Contemporary anesthesia guidelines now permit carbohydrate-containing clear liquids up to 2 hours before induction, reflecting safety data and benefits for patient comfort and insulin sensitivity.(9) Randomized and meta-analytic evidence indicates that preoperative carbohydrate loading can attenuate postoperative insulin resistance and improve early recovery metrics, with emerging analyses suggesting reductions in length of stay versus fasting or placebo.(10–12)

Beyond macronutrient timing, micronutrient status may modulate surgical resilience through redox, immune, and endothelial pathways. Trace elements and vitamins (e.g., selenium, zinc, copper, vitamins C/E) support endogenous antioxidant systems and wound healing, yet deficiencies are frequent in hospitalized and surgical populations and may worsen under operative stress.(13,14) Current guidance encourages early oral/enteral strategies tailored to surgical physiology, but there remains little randomized evidence on multi-nutrient perioperative formulations designed to target the acute immunometabolic response.(4)

Patient-centered outcomes such as postoperative nausea and vomiting (PONV) and pain remain highly prevalent and burdensome; risk stratification and multimodal prophylaxis are standard, yet residual morbidity is still common.(15,16) Preoperative affective state is another potentially modifiable factor: observational and meta-analytic data link higher preoperative anxiety with worse postoperative pain, sleep, delirium, and recovery outcomes, motivating trials that include emotional measures alongside biomedical endpoints.(17–19)

While ERAS-based carbohydrate regimens are supported, there is a paucity of pragmatic studies testing a multi-nutrient, generally recognized as safe (GRAS)-based medical food administered in the immediate preoperative window to address both glycemic control and perioperative symptoms while meeting modern fasting standards.(4,9) Given the associations between perioperative dysglycemia and adverse outcomes, and biologic plausibility for micronutrient support, a pilot trial focused on safety/tolerability with exploratory metabolic and patient-reported outcomes is warranted.

We conducted a single-center pilot randomized trial comparing a preoperative multi-nutrient medical food versus fasting control in adults undergoing elective surgery. The primary objective was to evaluate safety and tolerability within ERAS-consistent timing (including verification of gastric clearance). Secondary, exploratory objectives were to estimate effect sizes for pre, peri- and postoperative glycemic markers, electrolytes, cortisol, pain, PONV, and preoperative emotional state to inform a definitive efficacy trial. We hypothesized that the intervention would be safe and well tolerated and would generate favorable outcomes compared with control.

## Methods

This manuscript adheres to the CONSORT 2025 reporting guideline(20), along with the CONSORT extensions for pilot trials and non-pharmacologic treatments (NPT) (21,22).

### Study Design and Setting

We conducted a single-center, nonrandomized pilot clinical trial comparing a preoperative multi-nutrient medical food with fasting control, embedded in standard preoperative care pathways. The study took place at JSS Medical College, Mysuru, India, a community hospital. A priori, the trial focused on safety/tolerability with exploratory metabolic and patient-reported outcomes to estimate effect sizes for a subsequent efficacy trial. The study followed an *a priori* protocol established before enrollment but the trial was not registered.

### Participants

Eligible participants were adults (≥18 years) scheduled for elective abdominal, cardiac/thoracic, gynecological, or orthopedic surgery. Recruitment occurred preoperatively; written informed consent was obtained prior to study procedures. Exclusion criteria included kidney or liver disease, participation in a different clinical investigation that may affect the safety or performance of the investigation, being employees or family members of anyone involved in the investigation, and any other condition or treatment making a subject unsuitable for participation as judged by the Principal Investigator. Patient characteristics were recorded and are presented in Table 1.

**Table 1.** Patient characteristics.

|  |  | **Overall** | **Control** | **Intervention** | ***p*** |
| --- | --- | --- | --- | --- | --- |
| **n** |  | 67 | 34 | 33 |  |
| **age (mean (SD))** |  | 49.00 (15.05) | 50.53 (12.83) | 47.42 (17.10) | 0.403 |
| **sex (n, (%))** |  |  |  |  | 0.910 |
|  | F | 31 (46.3) | 15 (44.1) | 16 (48.5) |  |
|  | M | 36 (53.7) | 19 (55.9) | 17 (51.5) |  |
| **heart rate (mean (SD))** |  | 81.40 (7.92) | 82.85 (7.35) | 79.91 (8.32) | 0.129 |
| **systolic blood pressure (mean (SD))** |  | 131.99 (9.97) | 131.32 (11.78) | 132.67 (7.80) | 0.585 |
| **diastolic blood pressure (mean (SD))** |  | 82.76 (6.37) | 82.56 (6.81) | 82.97 (5.98) | 0.794 |

### Randomization and Masking

Participants were allocated to Treatment vs Control by the surgeons to match the two groups in age, sex, and surgery performed. The control arm was standard-of-care, which did not include a direct comparator of the medical food, so participant blinding was not feasible. To minimize performance bias in the absence of blinding, the product was administered by different staff than those who conducted the assessment of the patients. In addition, patients were given the same contact times and procedures throughout the study.

### Intervention

The investigational product was a commercially available multi-nutrient medical food, Vis Pre-Surgery™, comprising 15 micronutrients, trace minerals, and plant-based complex carbohydrates, all GRAS-designated (Table 2). The dosing schedule was two pre-operative administrations: one the evening before surgery and one ∼4 hours pre-induction, taken orally in addition to standard care (ERAS-consistent fasting). The comparator was fasting control with standard care.

**Table 2.** Nutrient Components and Rationale for the Perioperative Medical Food.

| **Ingredient** | **Class** | **Putative mechanism / rationale** | **Perioperative relevance** |
| --- | --- | --- | --- |
| **Trehalose** | Low-glycemic disaccharide | Protein/lipid stabilization; osmoprotection; may blunt rapid glycemic excursions | May support glycemic steadiness and cellular stress tolerance around induction |
| **Arabinogalactan (larch)** | Soluble fiber / prebiotic | Fermentable fiber leads to short-chain fatty acid (SCFA) production; supports commensal microbiota | SCFAs may modulate mucosal immunity and barrier function during surgical stress |
| **Magnesium** | Electrolyte / enzyme cofactor | Cofactor in >300 enzymes; neuromuscular and cardiac stability; glucose handling | Addresses common peri-op Mg fluctuations; supports rhythm/blood pressure and insulin signaling |
| **Pantothenic acid (B5)** | Water-soluble vitamin | Precursor of CoA; supports energy metabolism and steroidogenesis | Energy/recovery metabolism during catabolic stress |
| **Potassium** | Electrolyte | Membrane potentials; cardiac and muscle function | Maintains electrolyte balance under fluid shifts |
| **Vitamin C (ascorbic acid)** | Antioxidant vitamin | Redox cycling; collagen synthesis | Supports wound healing and antioxidant defenses |
| **Sodium** | Electrolyte | Extracellular volume/osmolality | Supports hemodynamic stability |
| **Monk fruit (mogrosides)** | Non-nutritive sweetener | Sweetness without glucose/fructose load; antioxidant polyphenols | Palatability while minimizing glycemic impact |
| **Zinc** | Trace element | Enzymatic cofactor; immune function; tissue repair | Supports innate/adaptive immunity and epithelial integrity |
| **Vitamin E (mixed tocopherols)** | Lipid-phase antioxidant | Protects membranes from lipid peroxidation | May limit oxidative injury during surgical stress |
| **Selenium** | Trace element | Cofactor for glutathione peroxidases/ thioredoxin reductases | Augments endogenous antioxidant systems |
| **Copper** | Trace element | Lysyl oxidase (collagen/elastin cross-linking); redox enzymes | Supports connective-tissue repair and erythropoiesis |
| **Vitamin D3 (cholecalciferol)** | Secosteroid vitamin | Calcium–phosphate homeostasis; immunomodulation | Bone health and immune tone during recovery |
| **Vitamin A (retinoids)** | Fat-soluble vitamin | Epithelial integrity; immune signaling | Supports barrier defenses and tissue repair |
| **Calcium** | Electrolyte / mineral | Excitation–contraction coupling; coagulation; bone | Supports neuromuscular function and hemostasis |
| *Natural berry flavor & other natural flavors* | Flavoring | Palatability / adherence | Improves acceptability of pre-op dosing |

### Outcomes

Primary safety/tolerability outcomes were:

1. Adverse events attributable to the study product

Secondary safety/tolerability outcomes were:

1. Postoperative nausea and vomiting (PONV) incidence
2. 30-day hospital readmission
3. Post-operative infections

Exploratory outcomes included:

1. Glycemic markers: Pre-op, peri-op, and post-op blood glucose (pre-op, intra/peri-op, 24 h, 48 h); HbA1c at pre-op and 90 days
2. Electrolytes: Post-op sodium, potassium, chloride, calcium, magnesium
3. Stress hormone: Cortisol (pre-op, post-op)
4. Patient-reported outcomes: Post-op pain (0–10) and pre-op emotional state (single-choice: Calm, Relaxed, Content, Tense, Worried, Upset).

### Sample Size and Statistical Analysis

This pilot study was designed to assess safety/tolerability and to generate effect-size estimates for a subsequent confirmatory trial. Per the a priori plan, analyses emphasized descriptive summaries and simple two-sided comparisons at α=0.05 without multiplicity adjustment. Standardized effect sizes accompany p-values to emphasize estimation over null-hypothesis testing.

The sample size per protocol was to be 200 participants. Enrollment was stopped early before complete enrollment due to frustrations around administrative issues at the study site. A third party auditor was sent in who verified the accuracy of the collected data and adherence to study protocol, but the decision was made to close enrollment at that time. The decision was unrelated to study outcomes and was made before data analysis.

All participants who received the assigned intervention and had outcome data were included in the full analysis set for exploratory efficacy. Continuous variables are summarized as mean (SD) and categorical variables as counts (%).

Between-group comparisons were specified as follows. For continuous endpoints (peri-/post-operative glucose, electrolytes, cortisol, and pain), the primary analysis used the Wilcoxon rank-sum (Mann–Whitney) test with two-sided α=0.05. To aid interpretation, we report the Hodges–Lehmann (HL) median difference from the rank-based analysis, alongside the standardized effect size Hedges’ g. We also present results from Welch’s two-sample *t*-test (mean difference) as a complementary analysis.

For HbA1c at 90 days, between-arm comparison followed the same nonparametric approach (Wilcoxon rank-sum). In addition, we fit an ANCOVA model with 90-day HbA1c as the dependent variable, study arm as the factor, and baseline HbA1c as a covariate to improve precision and to account for any baseline imbalance.

For binary endpoints (postoperative nausea/vomiting, readmission, infections, and positive pre-operative emotion), we used Fisher’s exact test (two-sided) and report risk ratios (RRs).

Analyses were performed in R(23) using base and contributed packages including tidyverse(24), janitor(25), tableone(26), broom(27), effectsize(28), epitools(29), and ggplot2(30).

### Ethics

This study was conducted in accordance with the Declaration of Helsinki and Good Clinical Practice (GCP). Ethical approval was obtained from the Independent Ethics Committee, JSS Medical College (IEC no. JSSMC/IEC/18-12-2023/20; approval date: December 18 2023).

All participants provided written, informed consent prior to any study procedures. Potential participants were given sufficient time to consider participation and ask questions. Signed consent forms and documentation of the consent discussion were maintained at the investigation site and made available for monitoring/auditing as required.

Study conduct, source documentation, and case report forms were monitored by study staff to verify protocol adherence, data accuracy, and protection of participants’ rights and welfare. An independent Data and Safety Monitoring Board (DSMB) was not convened because this was a small, single-center pilot study with a minimal-risk, food-based intervention and no anticipated safety concerns requiring formal interim monitoring. Routine internal monitoring was deemed sufficient by the ethics committee and investigators. Adverse events (AEs) were collected throughout participation and assessed for seriousness and relatedness and discussed with research staff to determine whether the AE was likely to be attributable to the intervention product.

Identifiable data were stored in secure, access-controlled systems at the site; de-identified data were used for analysis. Access to identifiable information was limited to authorized study personnel and monitors/auditors.

## Results

### Participant Flow and Baseline Characteristics

Sixty-seven adults undergoing elective surgery were enrolled and allocated to the medical food (Vis Pre-Surgery™, n=33) or no-treatment control (n=34). All participants completed the intervention dosing. A CONSORT-style flow diagram (Figure 1) summarizes screening, allocation, receipt of intervention, and inclusion in analyses, with reasons for any missing outcome data. Baseline characteristics (age, sex, blood pressure, heart rate) are summarized by arm in Table 1. The effects of all the outcome measures are displayed in Figure 2.

**Figure 1.**
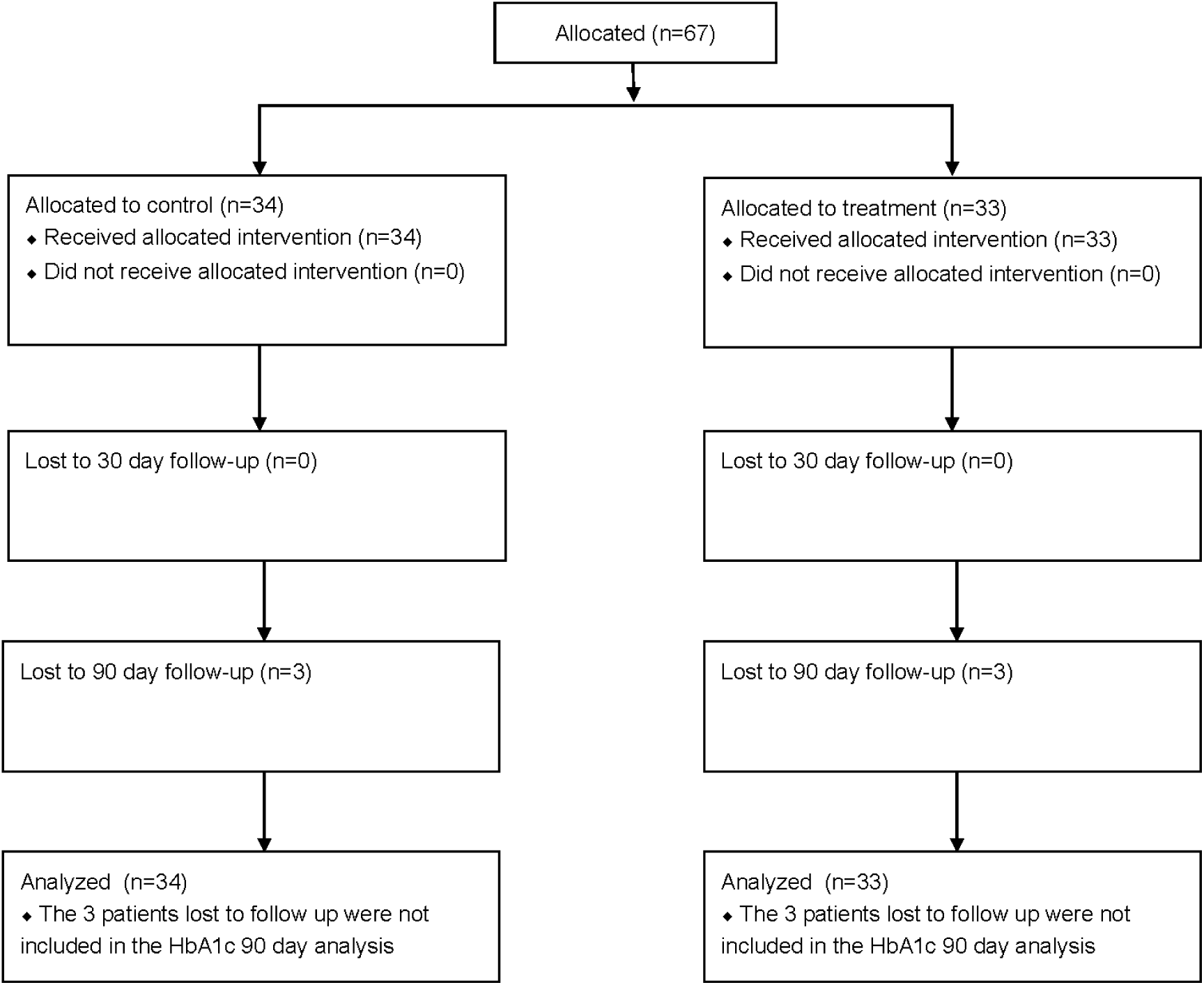
CONSORT Flow diagram

**Figure 2.**
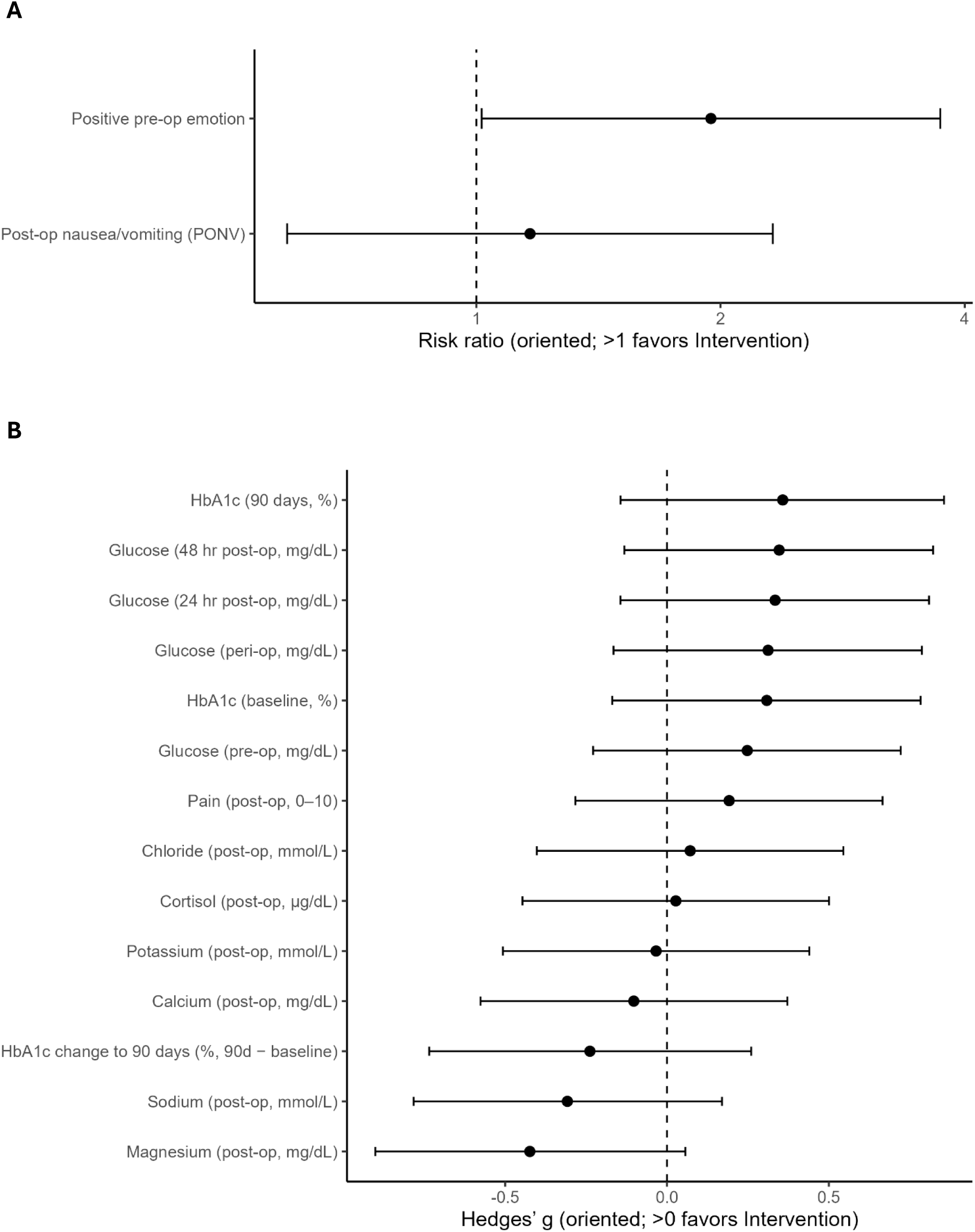
Between-group effects on binary and continuous endpoints A) Forest plot of risk ratios (RR) with 95% confidence intervals on a logarithmic x-axis. RRs are oriented so that values >1 favor the intervention; for PONV, the reciprocal RR is plotted to maintain this direction. The vertical dashed line denotes no effect (RR = 1). B) Forest plot showing standardized mean differences (Hedges’ g) with 95% confidence intervals for continuous outcomes. Effects are oriented so that positive values favor the intervention: for outcomes where lower values are desirable (glucose, HbA1c, pain), the sign is flipped. Vertical dashed line denotes no effect (g = 0).

### Primary and Secondary Outcomes (Safety and Tolerability)

No product-attributed adverse events occurred. Gastric clearance was confirmed within 2 hours after the pre-operative morning dose in all participants. There were no 30-day readmissions or post-operative infections in either arm. PONV rates were similar: 10/33 (30.3%) vs 12/34 (35.3%); RR 0.86, 95% CI 0.43–1.71, p=0.796.

### Exploratory Outcomes

#### Glycemic markers

Between-group differences in blood glucose were assessed with Wilcoxon rank-sum tests; Hodges–Lehmann (HL) median differences are reported as Intervention − Control. Across timepoints, estimates consistently favored lower glucose with the intervention.

Pre-operative glucose was lower in the intervention arm (means 126.06 vs 134.79 mg/dL; HL= −6.00, *p*=0.165; Hedges’ *g=* −0.25; Welch mean difference= −8.73, *p*=0.306). Peri-operative glucose showed a similar pattern (118.21 vs 129.29 mg/dL; HL= −8.00, *p*=0.103; *g=* −0.31; Welch= −11.08, *p*=0.199). At 24 hr post-op, the intervention again trended lower (121.45 vs 132.62 mg/dL; HL= −8.00, *p*=0.091; *g=* −0.33; Welch= −11.16, *p*=0.169). By 48 hr post-op, the between-arm difference reached statistical significance by the nonparametric test (125.03 vs 136.65 mg/dL; HL= −9.00, *p*=0.0297; Figure 3), while mean-based inference remained non-significant (*g=* −0.35; Welch= −11.62, *p*=0.154).

**Figure 3.**
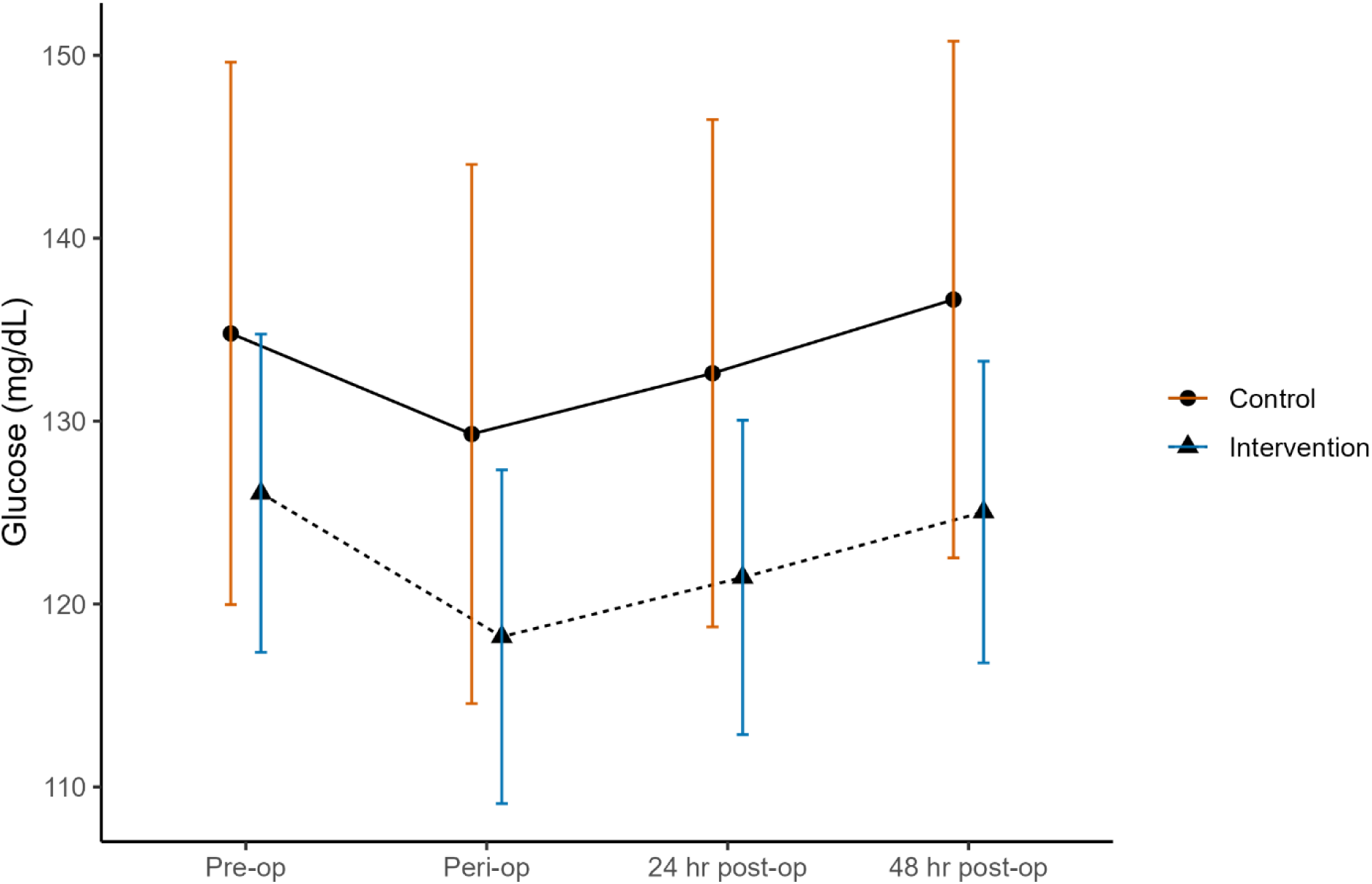
Peri-operative blood glucose over time by study arm (means with 95% CI) Mean blood glucose (mg/dL) at pre-op, peri-op, 24 hr post-op, and 48 hr post-op with 95% confidence interval error bars; lines connect timepoints within each arm.

#### HbA1c

Baseline HbA1c did not differ significantly between arms (means 5.23% vs 5.64%; HL= −0.10, *p*=0.628; *g=* −0.31; Welch= −0.41, *p*=0.204). At 90 days, unadjusted comparisons were likewise non-significant (5.00% vs 5.41%; HL= −0.07, *p*=0.845; *g=* −0.36; Welch= −0.41, *p*=0.160). Change through 90 days also did not differ (HL= −0.22, *p*=0.151; *g=* 0.24; Welch=0.09, *p*=0.350). In an ANCOVA adjusting 90-day HbA1c for baseline, the arm effect was null (*t*=−0.016; *p*=0.987), whereas baseline HbA1c strongly predicted 90-day values (*t*=31.579; *p*<0.001).

#### Electrolytes, cortisol, and pain

Post-operative electrolytes and cortisol showed no statistically significant between-arm differences by Wilcoxon rank-sum, although a difference in post-operative magnesium was trending, with small standardized and mean-based effects: sodium (HL= −1.00, *p*=0.427; *g=* −0.31; Welch= −1.52, *p*=0.211), potassium (HL= 0.10, *p*=0.642; *g=* −0.03; Welch= −0.02, *p*=0.889), chloride (HL= 0.00, *p*=0.781; *g=* 0.07; Welch= 0.35, *p*=0.769), calcium (HL= 0.00, *p*=0.826; *g=* 0.10; Welch= 0.08, *p*=0.670), magnesium (HL= −0.30, *p*=0.223; *g=* −0.42; Welch= −0.35, *p*=0.083), and cortisol (HL= 0.30, *p*=0.759; *g=* 0.03; Welch= 0.18, *p*=0.911). Mean post-operative pain scores were similar (5.33 vs 5.62 on a 0–10 scale; HL= 0.00, *p*=0.471; *g=* −0.19; Welch= −0.28, *p*=0.429).

#### Patient-reported pre-operative emotion

Participants in the intervention arm were twice as likely to endorse a positive pre-operative emotion (calm/relaxed/content) compared with control (17/33 vs 9/34); RR 1.95, *p*=0.046.

## Discussion

In this single-center pilot (n=67), a twice-dosed, carbohydrate-containing multi-nutrient drink given the evening before and ∼4 h before surgery was feasible, safe, and well-tolerated. Gastric clearance occurred within the targeted window for all participants, and there were no readmissions or infections within 30 days. Post-operative nausea and vomiting (PONV) did not differ significantly between arms; point estimates favored the intervention but with wide uncertainty, consistent with mixed evidence on PONV for pre-operative carbohydrate (CHO) drinks across procedures and co-interventions. At the same time, participants allocated to the medical food were more likely to endorse a positive pre-operative emotional state, a signal that aligns with trials showing CHO reduces pre-op hunger/thirst and improves well-being during the waiting period.(4,9,31)

Exploratory metabolic endpoints suggested modest glycemic benefits. Across the peri-/post-operative window, directionally lower glucose values were observed in the intervention arm at all four timepoints; the 48-hr difference reached statistical significance by the Hodges–Lehmann (HL) median-difference test but not by a mean-based Welch comparison. For HbA1c, between-arm differences were not statistically significant at baseline or 90 days, and an ANCOVA adjusting for baseline HbA1c showed no adjusted treatment effect while baseline strongly predicted follow-up. Using baseline-adjusted models here is methodologically preferable to analyzing change scores or raw follow-up means when baseline imbalances exist, because ANCOVA yields unbiased estimates and greater precision under common conditions.(32)

These findings are coherent with contemporary guidance and meta-analytic evidence. ASA’s 2023 practice update affirms that carbohydrate-containing clear liquids (with or without protein) are acceptable up to 2 h before elective anesthesia in healthy adults, supporting both the safety we observed and the practicality of a pre-induction dose.(9) ESPEN’s surgical nutrition guidance likewise endorses liberalized fasting and pre-op carbohydrate to reduce peri-operative discomfort and, particularly in major surgery, attenuate postoperative insulin resistance; importantly, across large meta-analyses, CHO loading reduces length of stay versus fasting but shows little to no effect on overall complication rates, mirroring the pattern in this pilot.(4)

On PONV specifically, the literature is mixed. A randomized trial in laparoscopic cholecystectomy found lower PONV 12–24 h after surgery with CHO versus fasting (33), but systematic syntheses generally report no clear overall reduction compared with water or placebo when antiemetic prophylaxis and procedure types vary, consistent with our neutral result. Procedure-specific protocols, dosing/timing, and multimodal antiemetic strategies likely explain some heterogeneity.(34)

Mechanistically, pre-induction carbohydrate may blunt counter-regulatory stress responses and mitigate insulin resistance, supporting more stable peri-operative glycemia; these effects have been demonstrated repeatedly in RCTs and reviews, even when complications are unchanged. Our multi-nutrient formulation also delivered micronutrients with plausible roles in redox balance and endothelial/immune support, but the present pilot was not designed to isolate nutrient-specific contributions.(12,35)

Finally, while not statistically significant, the trending pattern of lower magnesium in the intervention group (where magnesium was supplemented) is notable. Insulin signaling can drive magnesium from the extracellular space into cells; because preoperative carbohydrate loading improves perioperative insulin sensitivity, an intracellular shift could paradoxically lower serum magnesium shortly after surgery while cellular magnesium rises.(12,36,37) Thus, the directionally lower serum magnesium in the intervention arm is biologically plausible and does not necessarily indicate diminished magnesium status.

This study has important limitations. Allocation was not randomized or blinded, introducing risks of selection and performance bias despite efforts to standardize care. The comparator was usual care rather than an active CHO beverage commonly used in ERAS, so we cannot separate the value of a multi-nutrient composition from benefits achievable with standard carbohydrate loading. Small sample sizes at some endpoints limited precision (with multiple exploratory tests and no multiplicity control), and baseline differences in HbA1c complicate naive post-op contrasts, hence our emphasis on prespecified nonparametric tests and on baseline-adjusted models for confirmatory work. Although baseline HbA1c was lower in the intervention group, making unadjusted peri-operative glucose differences harder to interpret, it is nonetheless compelling that the only statistically significant difference occurred at 48 hours post-operatively. This aligns precisely with the period when stress-induced hyperglycemia typically peaks, reflecting maximal insulin resistance, counter-regulatory hormone activity, and metabolic stress. In a systematic analysis of postoperative glucose dynamics, the first 24–48 hours post-surgery was identified as a high-risk window for stress hyperglycemia and its complications(38–40). The emergence of between-arm separation specifically at 48 hours suggests the intervention may have blunted the expected hyperglycemic response above and beyond any influence from baseline HbA1c differences. This pattern strengthens the biological plausibility of the effect and highlights the 48-hour mark as a critical endpoint for confirmatory trials.

### Implications and next steps

These feasibility, safety, and signal-finding results justify a randomized, blinded (taste-matched) trial with a placebo or active CHO comparator, standardized multimodal PONV prophylaxis, and prespecified baseline-adjusted analyses for glycemic endpoints (e.g., ANCOVA or mixed models for repeated glucose/HbA1c). Based on effect sizes observed here, mean-based power calculations indicate that detecting clinically relevant differences in peri-/post-operative glucose would require on the order of ∼130–250 participants per arm (depending on timepoint) with two-sided α=0.05 and 80% power. Such a design would directly test whether a multi-nutrient approach offers incremental benefit over standard ERAS carbohydrate loading on metabolic control, PONV, patient-reported comfort, surgical throughput, and recovery, while remaining aligned with current fasting and nutrition guidelines.

## Funding

This was an industry-sponsored trial funded by TM Nutrition, LLC

## Conflicts of Interest

The authors declare the following competing interests: Thea Marx, ND is the Founder and Chief Executive Officer of TM Nutrition, LLC, which manufactures and sells the study product (Vis Pre-Surgery™) and funded this trial. TM Nutrition, LLC also provides partial salary support to Ben Zimmerman. All other authors declare no competing interests.

## Data Availability Statement

De-identified data supporting the findings and the analysis code are available upon reasonable request to the corresponding author, Thea Marx, ND.

## Author Contributions (CRediT)

**Conceptualization:** Thea Marx (TM).

**Methodology:** TM.

**Investigation:** TM.

**Formal analysis:** Benjamin Zimmerman (BZ).

**Data curation:** BZ.

**Writing—original draft:** BZ.

**Writing—review & editing:** BZ, Joshua Zvi Goldenberg (JZG), TM.

**Visualization:** BZ.

**Supervision:** TM, JZG.

**Project administration:** TM.

**Funding acquisition:** TM.

**Notes:** All authors meet ICMJE authorship criteria, had access to the data, contributed to manuscript revisions, and approved the final version.

## Notes

### Clinical Trial

NCT07359222

### Author Declarations

The Independent Ethics Committee of JSS Medical College gave ethical approval for this work (IEC no. JSSMC/IEC/18122023/20; approval date: December 18 2023).

